# Performance of Universal and Stratified Computer-Aided Detection Thresholds for Chest X-Ray-Based Tuberculosis Screening: A Cross-Sectional Diagnostic Accuracy Study

**DOI:** 10.1101/2025.04.09.25325458

**Authors:** Joowhan Sung, Peter James Kitonsa, Annet Nalutaaya, David Isooba, Susan Birabwa, Keneth Ndyabayunga, Rogers Okura, Jonathan Magezi, Deborah Nantale, Ivan Mugabi, Violet Nakiiza, David W Dowdy, Achilles Katamba, Emily A Kendall

## Abstract

**Background:** Computer-aided detection (CAD) software analyzes chest X-rays for features suggestive of tuberculosis (TB) and provides a numeric abnormality score. However, estimates of CAD accuracy for TB screening are hindered by the lack of confirmatory data among people with lower X-ray scores, including those without symptoms. Additionally, the appropriate X-ray score thresholds for obtaining further testing may vary according to population and client characteristics.

**Methods:** We screened for TB in Ugandan individuals aged ≥15 years using portable chest X-rays with CAD (qXR v3). Participants were offered screening regardless of their symptoms. Those with X-ray scores above a threshold of 0.1 (range, 0 – 1) were asked to provide sputum for Xpert Ultra testing. We estimated the diagnostic accuracy of CAD for detecting Xpert-positive TB when using the same threshold for all individuals (under different assumptions about TB prevalence among people with X-ray scores <0.1), and compared this estimate to age- and/or sex-stratified approaches.

**Findings:** Of 52,835 participants screened for TB using CAD, 8,949 (16.9%) had X-ray scores ≥0.1. Of 7,219 participants with valid Xpert Ultra results, 382 (5.3%) were Xpert-positive, including 81 with trace results. Assuming 0.1% of participants with X-ray scores <0.1 would have been Xpert-positive if tested, qXR had an estimated AUC of 0.92 (95% confidence interval 0.90-0.94) for Xpert-positive TB. Stratifying X-ray score thresholds according to age and sex improved accuracy; for example, at 96.1% specificity, estimated sensitivity was 75.0% for a universal threshold (of ≥0.65) versus 76.9% for thresholds stratified by age and sex (p=0.046).

**Interpretation:** The accuracy of CAD for TB screening among all screening participants, including those without symptoms or abnormal chest X-rays, is higher than previously estimated. Stratifying X-ray score thresholds based on client characteristics such as age and sex could further improve accuracy, enabling a more effective and personalized approach to TB screening.

**Funding:** National Institutes of Health

**Research in context:** *Evidence before this study:* The World Health Organization (WHO) has endorsed computer-aided detection (CAD) as a screening tool for tuberculosis (TB), but the appropriate X-ray score that triggers further diagnostic evaluation for tuberculosis (the “CAD threshold”) varies by population. The WHO recommends determining the appropriate CAD threshold for specific settings and population and considering unique thresholds for specific populations, including older age groups, among whom CAD may perform poorly. We performed a PubMed literature search for articles published until September 9, 2024, using the search terms “tuberculosis” AND (“computer-aided detection” OR “computer aided detection” OR “CAD” OR “computer-aided reading” OR “computer aided reading” OR “artificial intelligence”), which resulted in 704 articles. Among them, we identified studies that evaluated the performance of CAD for tuberculosis screening and additionally reviewed relevant references. Most prior studies reported area under the curves (AUC) ranging from 0.76 to 0.88 but limited their evaluations to individuals with symptoms or abnormal chest X-rays. Some prior studies identified subgroups (including older individuals and people with prior TB) among whom CAD had lower-than-average AUCs, and authors discussed how the prevalence of such characteristics could affect the optimal value of a population-wide CAD threshold; however, none estimated the accuracy that could be gained with adjusting CAD thresholds between individuals based on personal characteristics.

*Added value of this study:* In this study, all consenting individuals in a high-prevalence setting were offered chest X-ray screening, regardless of symptoms, if they were ≥15 years old, not pregnant, and not on TB treatment. A very low X-ray score cutoff (qXR v3 TB score of 0.1 on a 0-1 scale) was used to select individuals for confirmatory sputum molecular testing, enabling the detection of radiographically mild forms of TB and facilitating comparisons of diagnostic accuracy at different CAD thresholds. To assess CAD performance among all X-ray screening participants capable of providing sputum, a TB prevalence of 0.1% was assumed for individuals with X-ray scores <0.1 who were not offered sputum testing. Using this symptom-neutral evaluation of CAD with expansive criteria for bacteriologic testing, we estimated an AUC of 0.92, and we found that the qXR v3 threshold needed to decrease to under 0.1 to meet the WHO target product profile goal of ≥90% sensitivity and ≥70% specificity. CAD performance decreased when a higher prevalence of TB was assumed among people with X-ray score <0.1. Compared to using the same thresholds for all participants, adjusting CAD thresholds by age and sex strata resulted in a 1 to 2% increase in sensitivity without affecting specificity.

*Implications of all the available evidence:* To obtain high sensitivity with CAD screening in high-prevalence settings, low score thresholds may be needed. However, countries with a high burden of TB often do not have sufficient resources to test all individuals above a low threshold. In such settings, adjusting CAD thresholds based on individual characteristics associated with TB prevalence (e.g., male sex) and those associated with false-positive X-ray results (e.g., old age) can potentially improve the efficiency of TB screening programs.

## Introduction

Over ten million people are estimated to develop tuberculosis (TB) each year, of whom more than 3 million are never reported to public health authorities.^1^ Improved case finding strategies are urgently needed to reduce the global burden of TB.^2^ Chest X-ray (CXR) is a useful tool for TB screening, with higher sensitivity than symptom-based screening and potential for high throughput at low cost.^3^ Computer-aided detection (CAD) systems, which use artificial intelligence (AI) to analyze chest X-rays, have recently emerged as a promising tool for scaling up rapid CXR-based TB screening.

An important point of uncertainty is the most appropriate threshold at which to refer screening participants for further evaluation. CAD products typically generate a score (“X-ray score”) that correlates with the probability of pulmonary TB. The World Health Organization (WHO) recommends CAD calibration studies^4^ to determine the appropriate X-ray score threshold (“CAD threshold”) for each specific population and context. However, most existing studies have either evaluated the diagnostic accuracy of CAD among symptomatic individuals in clinical triage settings^5-10^ or offered sputum testing to symptom-negative people only if they had TB-suggestive X-rays.^11-15^ As a result, very limited data exist on the accuracy of CAD – and the optimal CAD threshold for further evaluation – among people with mildly abnormal chest radiographs and no known symptoms. These data are important given the likely contribution of asymptomatic TB to transmission of *M. tuberculosis* in communities.^16^

In developing optimal CAD thresholds for community-based screening, ancillary data – such as age and sex – may be particularly important to consider. Most existing evaluations of CAD for TB screening have used a single threshold for all participants,^17-20^ thereby ignoring known associations of TB risk with sex (i.e., higher prevalence in men)^21,22^ and age (i.e., higher probability of X-ray abnormalities representing non-TB conditions in older individuals).^23^ Therefore, tailoring X-ray score cutoffs based on age and sex may improve performance. We therefore analyzed results from an ongoing TB case-finding study in Uganda to evaluate the diagnostic accuracy of CAD among screening participants and assess the impact of stratifying CAD thresholds according to participant demographics.

## Methods

### Systematic TB screening with portable X-ray and CAD

We conducted community-based TB screening using portable digital chest X-ray with CAD as part of an ongoing cluster-randomized trial in Uganda (CHASE-TB, Clinicaltrials.gov: NCT05285202). Participants were recruited to undergo screening in testing tents set up either near district-level health facilities (“facility-based” sites) or in areas with high traffic, such as transit hubs or markets, in neighborhoods and villages believed to have high TB prevalence (“community-based” sites), in peri-urban and rural areas surrounding Kampala. At facility-based sites, recruitment was not limited to patients seeking care at the facilities, but also included companions, staff, and passersby. At community-based sites, participants were recruited by interacting with anyone passing by, and by visiting nearby homes and shops when enrollment slowed. At both sites, all adults or adolescents ≥ 15 years old and not on active TB treatment were eligible for the study, regardless of symptoms. All consenting participants completed a standard questionnaire that included demographics, smoking history (added five months after study initiation), known TB exposures, 30-day TB symptom history, TB treatment history, and HIV status. All non-pregnant participants were offered screening with digital chest X-ray using a portable X-ray device. Chest radiographs were then read in real-time by CAD software (qXR v3.0, [Qure.ai, India]) independently of all clinical data. Participants whose X-rays were assigned qXR TB scores (“X-ray scores”; range: 0-1) higher than the prevailing threshold were asked to provide expectorated sputum, which was sent for Xpert MTB/RIF Ultra (“Xpert”) testing at a local health facility (without accompanying clinical information or X-ray results). X-ray scores are intended to discriminate TB status but are not directly interpretable as probabilities. The CAD threshold for Xpert testing was initially set to 0.5 and was adjusted to 0.2 after one month and to 0.1 after four additional months, reflecting the distribution of X-ray scores and desire to utilize available testing capacity for research purposes. The present analysis retrospectively considers data from all participants from both facility-based and community-based sites who were screened from June 1, 2022 (study start) through March 31, 2024 (see **appendix p 3** for power calculation).

### Statistical analysis

Participant characteristics were summarized as medians with interquartile ranges [IQRs], or as percentages for categorial variables, and were compared across groups using t-tests and chi-squared tests.

We estimated the performance (sensitivity, specificity, and area under the curve [AUC]) of CAD for detecting Xpert-positive TB among non-pregnant individuals who could provide an expectorated sputum sample. Because participants with X-ray scores under 0.1 were not asked for sputum, our primary analysis assumed that 0.1% of participants with X-ray scores <0.1 would be Xpert-positive (an estimate supported by analysis of CAD data from a study of universal Xpert Ultra screening in Uganda, **appendix pp 4-6**), with sensitivity analyses assuming Xpert-positive proportions ranging from 0.05% (half of our primary assumption as the lower limit) to 0.3% (the estimated national prevalence of Xpert-positive TB in Uganda^25^). For participants with an X-ray score <0.1, we assumed that the proportion who would successfully provide a sputum sample was similar to that of participants with scores between 0.1 and 0.19 (**appendix p 6**), and we assigned sputum production and Xpert status randomly.

We derived age- and/or sex-stratified CAD thresholds under the principle that, to maximize the impact of screening under constrained confirmatory testing capacity, Xpert tests should be offered with a similar minimum pre-test probability in all participant subgroups. To derive age- and/or sex-stratified CAD thresholds, we first fitted shape-constrained (monotonically increasing) generalized additive models^26^ for each subgroup, using Xpert result as the outcome and X-ray score as the explanatory variable. We then identified, for each age and/or sex subgroup, the score at which the prevalence of Xpert positivity was estimated to be closest to 2% (and separately to 1%, as a sensitivity analysis), corresponding to a resource threshold of 50 (and 100) Xpert tests required to produce one positive result. We took these scores as the age- and/or sex-stratified CAD thresholds for each age/sex subgroup, and we estimated the overall sensitivity and specificity of CAD when those age- and/or sex-stratified CAD thresholds were used. Then, to compare the performance of a universal strategy against age- and/or sex-stratified approaches, we identified the universal CAD threshold that would match the specificity of each set of stratified thresholds, representing a fixed capacity for confirmatory testing of people who do not have TB. We compared sensitivities between the universal and stratified approaches, to estimate the potential sensitivity gains achievable by stratifying thresholds under constrained confirmatory testing resources. We considered trace-positive Xpert Ultra results as positive in our primary analyses,^27^ but we also performed sensitivity analyses considering them negative.

Because participants with X-ray scores between 0.1 and 0.49 were not asked for sputum during the first five months of the 22-month study, we limited CAD threshold selection and accuracy evaluation to participants enrolled after the CAD threshold was lowered to 0.1. We then conducted a sensitivity analysis that included data from participants screened before the threshold change, using bootstrapping to recreate screening populations of the same size and X-ray score distribution as the full study population but with complete Xpert information (**appendix p 8**). Outcomes were estimated using the same set of thresholds selected in the primary analysis based on the more limited dataset, along with corresponding uncertainty. Statistical significance was defined as two-sided p<0.05. Analyses were conducted using Stata version 16.1 and R version 4.3.2.

### Ethics statement

The study was approved by the Institutional Review Boards at the Johns Hopkins University School of Medicine and Makerere University School of Public Health. Oral informed consent (or assent with parental consent) was obtained from all study participants.

## Results

A total of 54,840 individuals aged ≥ 15 years were assessed for study eligibility. Seventy-seven were on TB treatment, 12 did not consent, 1,374 were pregnant and offered sputum testing without X-ray, and 542 eligible individuals did not have an X-ray result documented. Therefore, a total of 52,835 participants were screened for TB using AI-interpreted digital X-rays with valid X-ray scores, including 45,758 screened after the CAD threshold was lowered to 0.1. Of the 52,835 screened participants, the median age was 38 years, 44.6% were male, 6.6% reported known HIV infection, and 1.4% reported a prior history of TB treatment (**Table 1**). About one-third (31.9%) reported cough within the past 30 days.

**Table 1:**
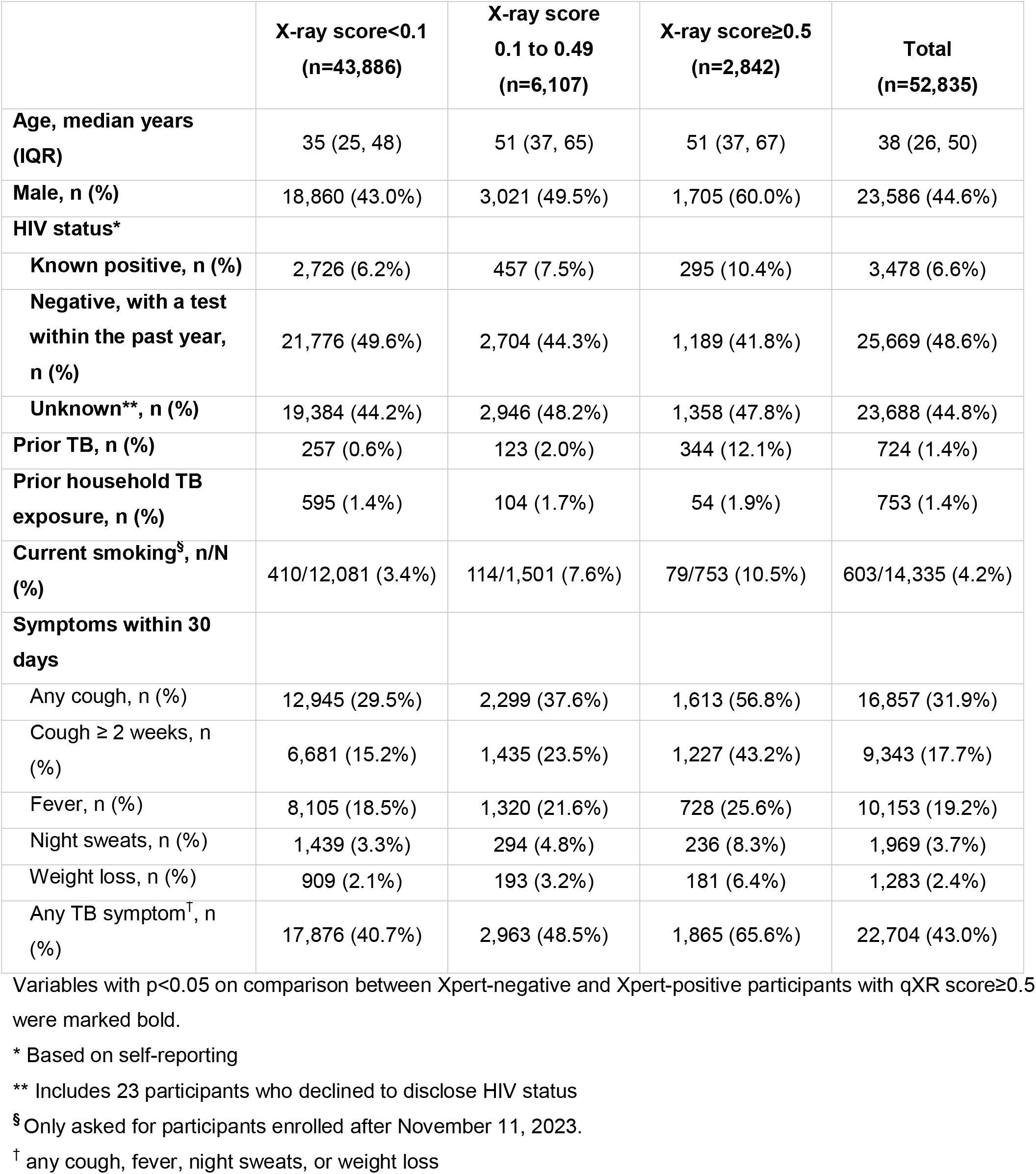
Characteristics of participants aged 15 years and older who received digital chest X-ray based TB screening with computer-aided detection software (qXR)

Of 52,835 screened participants, 43,886 (83.1%) had an X-ray score <0.1, 6,107 (11.6%) had a score between 0.1 and 0.49, 1,616 (3.1%) had a score between 0.5 and 0.89, and 1,226 (2.3%) had a score ≥0.9 (**Figure 1a**). A total of 8,038 (15.2%) were offered sputum testing, of whom 7,239 (90.1%) provided sputum and 7,219 (89.8%) had valid Xpert Ultra results. Of valid Xpert results, 301 (4.2%) were positive at a level greater than trace, and an additional 81 (1.1%) were trace-positive. Younger age, male sex, TB symptoms, and prior TB treatment were associated with positive Xpert results (including trace) (**appendix p 9**), including among individuals with X-ray scores between 0.1 and 0.5 (**appendix p 10**).

**Figure 1.**
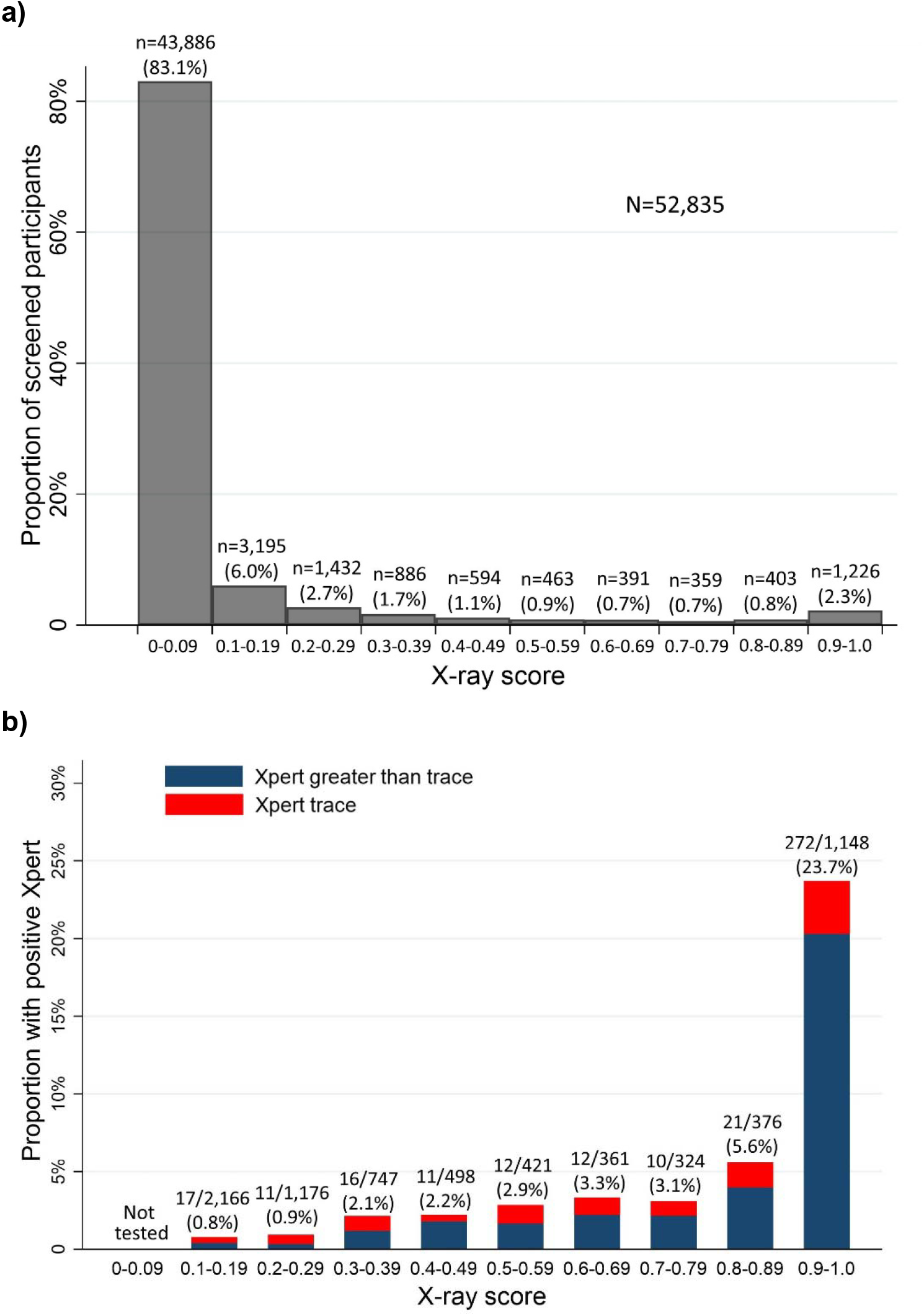
X-ray and Xpert results of community-screened adults (≥15 years) in Uganda. Panel (**a**) shows the distribution of qXR TB scores among study participants who received X-ray-based TB screening in Uganda with computer-aided detection software (qXR). Each bar height represents the proportion of participants whose X-ray scores were in the corresponding range. Panel (**b**) illustrates the proportions of participants whose sputum Xpert MTB/RIF Ultra results were positive within each X-ray score increment. Bar coloring differentiates the proportions of positive Ultra results that were greater than trace (blue) or trace (red).

**Figure 1b** illustrates the relationship between X-ray scores and Xpert results. Of participants with X-ray scores between 0.1 and 0.2, 0.8% (17 out of 2,166) had positive (including trace-positive) Xpert results. The prevalence of Xpert positivity increased to 2.5% among participants with scores between 0.4 and 0.59 and reached 23.7% (272 out of 1,148) among those with scores of 0.9 or higher (**Table 2)**. The proportion of study participants found to have Xpert-positive sputum was higher for men (1.2%) than women (0.3%), and was similar between age groups (0.8% if ≥40 years vs. 0.7% if <40 years). However, older participants had a higher prevalence of X-ray abnormalities detected by CAD (X-ray score ≥0.1 in 26.0% vs. 9.0%) (**appendix p 11**).

**Table 2:** Proportion of participants with positive sputum Xpert Ultra results, by sex, age group, and X-ray score. Shaded cells indicate subgroups with Xpert positive prevalence ≥1% (light gray) or ≥2% (dark gray).

|  | Proportion of participants with positive* Xpert results, among those tested |  |  |  |  |
| --- | --- | --- | --- | --- | --- |
| X-ray score | Male | Female | < 40 years old | $\geq 40$ years old | All |
| **0.1–0.19** | 14/1042 (1.3%) | 3/1124 (0.3%) | 7/671 (1.0%) | 10/1495 (0.7%) | 17/2166 (0.8%) |
| **0.2–0.29** | 8/590 (1.4%) | 3/586 (0.5%) | 4/299 (1.3%) | 7/877 (0.8%) | 11/1176 (0.8%) |
| **0.3–0.39** | 9/367 (2.5%) | 7/380 (1.8%) | 9/173 (5.2%) | 7/574 (1.2%) | 16/747 (2.1%) |
| **0.4–0.49** | 8/250 (3.2%) | 3/248 (1.2%) | 3/106 (2.8%) | 8/392 (2.0%) | 11/498 (2.2%) |
| **0.5–0.59** | 9/240 (3.8%) | 3/181 (1.7%) | 3/87 (3.5%) | 9/334 (2.7%) | 12/421 (2.9%) |
| **0.6–0.69** | 9/198 (4.6%) | 3/163 (1.8%) | 4/75 (5.3%) | 8/286 (2.8%) | 12/361 (3.3%) |
| **0.7–0.79** | 9/168 (5.4%) | 1/156 (0.6%) | 7/89 (7.9%) | 3/235 (1.3%) | 10/324 (3.1%) |
| **0.8–0.89** | 14/198 (7.1%) | 7/178 (3.9%) | 6/97 (6.2%) | 15/279 (5.4%) | 21/376 (5.6%) |
| **0.9–1.0** | 209/789 (26.5%) | 63/359 (17.6%) | 150/402 (37.3%) | 122/746 (16.4%) | 272/1,148 (23.7%) |
| **Any X-ray score $\geq 0.1$** | 289/3,843 (7.5%) | 93/3,376 (2.8%) | 193/1,999 (9.7%) | 189/5,220 (3.6%) | 382/7,219 (5.3%) |

\* Including trace-positive

Among 45,758 (out of 54,840, 83.4%) participants screened after the CAD threshold was lowered to 0.1, the estimated X-ray scores corresponding to a 1% or 2% probability of Xpert positivity were: 0.11 or 0.47 for men aged 15-39, 0.25 or 0.53 for men aged ≥40, 0.28 or 0.52 for women aged 15-39, and 0.44 or 0.89 for women aged ≥40. Among all participants with positive Xpert results, semiquantitative Xpert results were weakly correlated with X-ray scores (Spearman’s correlation coefficient=0.28) **(appendix p 12)**.

Assuming that 0.1% of people with X-ray scores below 0.1 would test positive on sputum Xpert, community-based screening using CAD had an estimated AUC of 0.92 (95% confidence interval [CI] 0.90-0.94) for Xpert-positive TB **(Figure 2)**. Under this assumption, the manufacturer-recommended universal threshold of 0.5 had an estimated sensitivity of 79.1% (95% CI: 74.3-83.2) and specificity of 94.8% (94.6-95.0) (**appendix p 13)**. Lowering the threshold to 0.1 improved sensitivity to 89.9% (95% CI 86.1-92.7) while lowering specificity to 83.6% (95% CI 83.2-83.9). Sensitivity varied by screening location but remained above 85% at a threshold of 0.1 across all eight regions (**appendix p 14**). When we assumed a 0.3% prevalence of Xpert-positive TB among people with X-ray score <0.1, the AUC decreased to 0.83 (95% CI 0.81-0.86), sensitivity fell to 65.6% (95% CI 60.7-70.2) at a threshold of 0.5, or 74.5% (95% CI 69.9-78.7) at a threshold of 0.1, and specificity remained similar (94.8% [94.6-95.0] at a threshold of 0.5 and 83.5 [83.2-83.9] at a threshold of 0.1). In the sensitivity analysis incorporating data from participants enrolled early in the study, the AUC remained similar at 0.93 (95% CI 0.91-0.94), assuming 0.1% prevalence among those with X-ray scores <0.1 (**appendix p 15**).

**Figure 2.**
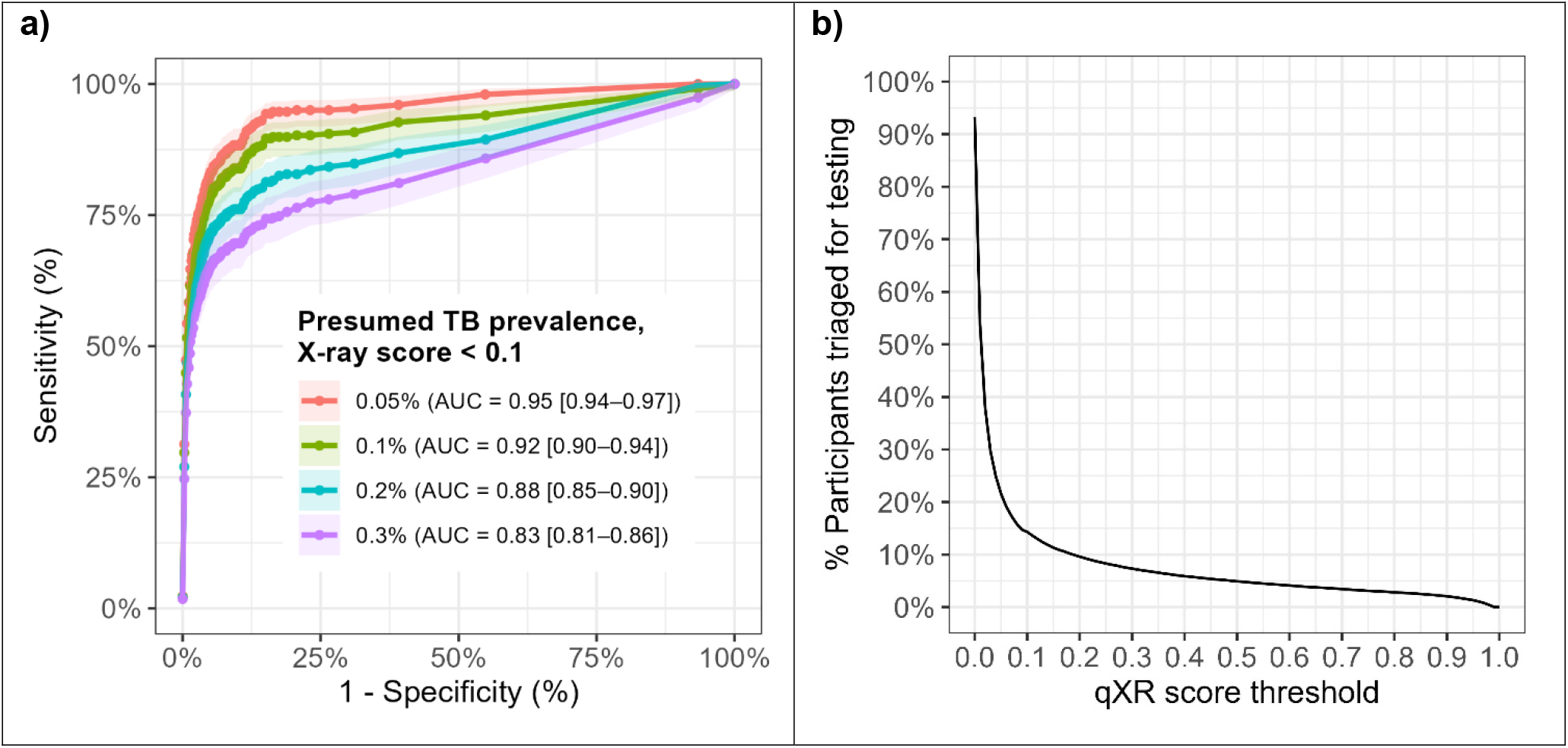
Diagnostic performance of CAD-based TB screening in Ugandan communities. **Panel A** shows receiver operating characteristic (ROC) curves illustrating diagnostic performance of computer-aided detection software (qXR version 3) for sputum Xpert-positive TB during community-based TB screening in Uganda. The colors of the ROC curves represent different assumptions about the prevalence of TB among people with an X-ray score <0.1 who did not qualify for sputum Xpert testing (red line: 0.05% prevalence; green line: 0.1% prevalence; blue line: 0.2% prevalence; purple line: 0.3% prevalence). Uncertainty is shown as 95% confidence intervals on the ROC curves and the AUC estimates. **Panel B** shows, for each possible X-ray score threshold between 0.0 and 1.0, the proportion of participants who had X-ray scores greater than or equal to that value and would thus qualify for TB testing at that threshold.

Compared to a universal threshold with matching specificity, age- and sex-stratified thresholds, set at the scores corresponding to estimated 2% Xpert positivity in all subgroups, had higher sensitivity (76.9% versus 75.0%, p=0.046) (**Table 3**). The sensitivity differences between age- and sex-stratified versus universal thresholds were similar when stratified thresholds were selected to result in 1% Xpert positivity (85.1% versus 83.5%, p=0.182) or when higher or lower Xpert positivity were assumed for people with qXR scores <0.1 (81.0% versus 79.0% assuming 0.05% prevalence, or 63.8% versus 62.2% assuming 0.3% prevalence). Age- and sex-stratified thresholds also resulted in higher sensitivities when trace-positive results were considered as negative (79.1% versus 77.6%, p=0.221) or when all eligible participants, including those who screened early in the study, were accounted for (73.4% versus 72.4%, p=0.210) (**appendix pp 16-19**). Stratification by sex alone resulted in a smaller gain in sensitivity at higher thresholds (75.9% versus 75.0%, p = 0.546 or 87.0% versus 87.0% at lower thresholds, p=1.000), while stratification by age alone did not consistently improve sensitivity (76.6% versus 77.2%, p=0.480 or 82.9% versus 82.3%, p=0.683) (**appendix p 20**).

**Table 3:** Sensitivity and specificity of age- and sex-stratified qXR thresholds for detecting Xpert-positive tuberculosis, compared with universal thresholds of matching specificity.

| Assumed TB prevalence when X-ray score <0.1 | Fixed-specificity comparison, higher sensitivities |  | Fixed-specificity comparison, lower sensitivities |  |
| --- | --- | --- | --- | --- |
| | Age- and sex-stratified threshold* (15-39y M: $\geq 0.11$; $\geq 40y$ M: $\geq 0.25$; 15-39y F: $\geq 0.28$; $\geq 40y$ F: 0.44) | Universal threshold ($\geq 0.26$ for all<sup>†</sup>) | Age- and sex-stratified threshold* (15-39y M: $\geq 0.47$; $\geq 40y$ M: $\geq 0.53$; 15-39y F: $\geq 0.52$; $\geq 40y$ F: 0.89) | Universal threshold ($\geq 0.65$ for all<sup>†</sup>) |
|  | Sensitivity Estimate (95% CI) |  |  |  |
| 0.05% | 89.7<br>(85.7-92.6) | 88.0<br>(83.8-91.2) | 81.0<br>(76.2-85.0) | 79.0<br>(74.0-83.2) |
| 0.1% | 85.1<br>(80.8-88.6) | 83.5<br>(79.1-87.2) | 76.9<br>(71.9-81.2) | 75.0<br>(69.9-79.5) |
| 0.2% | 77.3<br>(72.6-81.4) | 75.9<br>(71.1-80.1) | 69.8<br>(64.8-74.4) | 68.1<br>(63.0-72.8) |
| 0.3% | 70.6<br>(65.8-75.0) | 69.3<br>(64.5-73.7) | 63.8<br>(58.8-68.4) | 62.2<br>(57.2-66.9) |
|  | Specificity Estimate (95% CI) |  |  |  |
| 0.05 to 0.3%<sup>‡</sup> | 91.0<br>(90.7-91.3) | 91.0<br>(90.7-91.3) | 96.1<br>(95.9-96.3) | 96.1<br>(95.9-96.3) |

\* Correspond to the estimated scores at which approximately 1% (left columns) or 2% (right columns) of participants in each subgroup would test positive.
<sup>†</sup> Chosen to match the 91.0% (left columns) or 96.1% (right columns) aggregate specificity of age- and sex-stratified thresholds.
<sup>‡</sup> Specificities remained the same when TB prevalence among individuals with X-ray scores <0.1 was assumed to be 0.05%, 0.1%, 0.2%, or 0.3%.

## Discussion

In this evaluation of an active case-finding program in Uganda, the diagnostic accuracy of CAD, particularly specificity, was higher than in prior studies^11,13-15^ that restricted evaluation of CAD performance to screening participants with TB symptoms or abnormal X-rays. Under the assumption that no more than 0.1% of people with normal or near-normal X-rays (X-ray score <0.1) have Xpert-positive TB, the AUC for CAD in detecting Xpert-positive TB in all screening participants able to provide sputum (regardless of their symptoms or X-ray results) was 0.92, and it would be possible for a threshold near 0.1 to meet the WHO target product profile (TPP) goal^28^ of ≥90% sensitivity and ≥70% specificity. This accuracy can potentially be further improved by stratifying the CAD threshold by participant age and sex. These results speak to the utility of CAD in the population-based screening context and argue for further investigation of liberal CAD thresholds that are stratified by readily measured participant characteristics.

In this population, about 5% of participants had an X-ray score over the manufacturer-recommended threshold of 0.5 and these had the highest probability of a positive Xpert result. However, the additional 12% of participants with an X-ray score between 0.1 and 0.49 still had a >1% risk of having Xpert-positive TB, and excluding them from confirmatory testing limited sensitivity to 79%. Therefore, in the setting of community-based screening, lower CAD thresholds may be necessary to achieve desired sensitivity. Our finding of the wide radiographic spectrum of TB in communities is aligned with a recent CAD analysis of South African prevalence survey participants,^15^ where an CAD threshold needed to be lowered to 0.18 to achieve 90% sensitivity. As lowering the CAD threshold can strain limited resources in most high TB-burden areas, the costs and benefits of more sensitive versus more stringent screening strategies should be weighed when setting thresholds. Formal decision analysis, informed by cost-effectiveness analysis, could help set appropriate thresholds for different populations.

We also demonstrated that tailoring CAD thresholds according to easily identifiable individual characteristics, such as age and sex, has potential to improve the accuracy of TB screening. Among all study participants, the prevalence of TB was four times higher in male than in female participants. As a result, male participants with X-ray scores ≥0.52 had a comparable TB risk as females with X-ray scores ≥0.81. In contrast, while the prevalence of TB was similar between all participants under and over 40 years old, older individuals were almost three times more likely to have abnormal chest imaging (i.e., X-ray score ≥0.1) compared to younger individuals, likely due to higher prevalence of other lung conditions (including unrecognized prior TB) among older adults. Consequently, among individuals who qualified for Xpert testing with abnormal CXR, younger individuals were three times more likely to test positive on Xpert than older individuals, and individuals under 40 years old with an X-ray score ≥0.47 had a comparable risk for TB as those 40 years and older with an X-ray score ≥0.62. Therefore, in settings with limited resources, the use of age- and sex-stratified thresholds has the potential to increase the number of TB cases detected within the existing confirmatory testing capacity.

Our finding represents a promising opportunity to offer personalized screening in the context of active case-finding. In clinical settings, multiple factors, such as age, sex, HIV status, exposure history, symptoms, and chest imaging, are all incorporated into the decision-making process when deciding to test for TB. In mass-screening settings, this level of assessment is often not feasible, and simple criteria (such as presence of TB symptoms or abnormal CXR) are frequently used instead to select individuals for further testing. However, the emergence of CAD technology – which provides quantitative outputs that correlate with probability of Xpert positivity – now make it possible to incorporate additional TB risk factors into the screening algorithm. While the increase in sensitivity using age/sex stratification was modest (1-2%), this could nonetheless be valuable in resource-limited settings where confirmatory testing cannot be offered to all individuals who have X-ray scores above a low threshold. Moreover, implementing individualized thresholds using very simple characteristics such as age and sex will likely incur minimal additional costs. If validated prospectively and in other populations, setting stratified thresholds, including lowering thresholds for high-risk individuals, should be considered.

Lastly, we note that our estimates of specificity at a given sensitivity in the systematic TB screening context are high relative to some other studies, due to a difference in methodology. Even at the lowest CAD threshold for which we collected bacteriologic data, 0.1, our estimate of specificity remained at 83.6% (well above the TPP goal of ≥70%). Three prior studies^13,15,18^ that evaluated qXR v3 reported much lower specificities, ranging from under 50% to 62% for X-ray score thresholds providing 90% sensitivity. Those lower estimates resulted from limiting evaluations to participants with valid Xpert results, and thus to people with symptoms or abnormal CXR. By contrast, by using a low CAD threshold and estimating the Xpert prevalence even below that threshold, we estimated accuracy among an entire mass-screening population, including the large segment with negative symptom screens and normal X-rays for whom TB testing is not typically performed. This inclusion of all screening-eligible individuals in accuracy estimates increases the number of true negatives (i.e., no TB, with low X-ray scores) and results in higher specificities of CAD, as reported by studies^29,30^ including ours, that included individuals with normal X-rays in CAD accuracy estimation.

Our study has certain limitations. Although we used a low CAD threshold, we did not perform Xpert testing for individuals with X-ray scores < 0.1. Since about 80% of our participants fell into this category, our estimates of sensitivity depend considerably on the prevalence of TB among individuals with X ray scores <0.1. Our findings regarding the AUC of qXR and the added value of stratified thresholds were reasonably robust to sensitivity analysis around the prevalence of TB in this population. Secondly, we evaluated accuracy relative to a pragmatic reference standard of expectorated sputum Xpert (without sputum induction or culture). While accuracy relative to a more comprehensive reference standard is therefore unknown, expectorated sputum Xpert confirmatory testing is common practice in systematic screening, and our estimates therefore reflect the ability of CAD to lead to detection of sputum Xpert-positive tuberculosis. Additionally, our analysis did not include pregnant participants, nor participants who failed to provide expectorated sputum (1.6% of all participants or 10% of those with X-ray score ≥0.1). Although we evaluated stratification only by age and/or sex due to the ready availability of these variables, further work should explore stratification by other characteristics, including HIV status, which was unknown for about half of our participants, and history of prior TB. In addition, we evaluated only one CAD software, and we did not have human readers against whom to compare CAD readings. Thus, we are unable to identify specific radiographic features associated with TB at low X-ray scores. Finally, the accuracy of stratified thresholds depended on the Xpert status of a relatively small number of people with X-ray scores between the two peaks of a bimodal X-ray score distribution and may be sensitive to changes in CAD performance. Our findings therefore warrant validation, including use of other CAD software solutions and other populations, as well as investigation into potential challenges in implementing stratified thresholds. If resources permit, evaluating TB prevalence among individuals with low X-ray scores would help further determine the utility of chest X-ray and CAD in TB screening, as CAD performance depends heavily on the prevalence of TB among the large number of participants with X-ray scores under the confirmatory testing threshold.

In summary, we screened over 50,000 individuals for TB in Ugandan communities using portable chest X-ray with CAD and found that, while CAD has overall high accuracy for Xpert-positive TB in this community-screening context, individuals with X-ray scores between 0.1 and 0.5 were still at elevated risk for TB. We showed that using age- and sex-stratified CAD thresholds can improve the accuracy of CAD for TB screening. While our findings need validation, adopting lower CAD thresholds should be considered where feasible, and in high-burden settings with limited confirmatory testing capacity, stratifying thresholds by age and sex may offer a more effective and personalized approach to TB screening.

## Data Availability

The de-identified dataset of participant demographics, X-ray scores, and Xpert results used for this study and a data dictionary will be available upon a reasonable request. Data sharing will be limited to non-commercial research use only. Requests should include a proposal outlining the intended use and methodology and will be subject to review and approval. Requests can be directed to.

## Contributors

JS conceptualized the research question, developed the analytic plan, directly accessed and verified the underlying data, performed the data analysis, and wrote the original draft of the manuscript. PJK coordinated field data collection activities in Uganda and contributed to manuscript preparation. AN contributed to data management and data curation. SB, KN, RO, JM, DN, IM, and VN enrolled participants and collected data in Uganda. DWD and EAK conceptualized the study, acquired funding and resources, supervised the data collection and development of the analytic plan, and critically revised the manuscript. EAK supervised the analysis and directly accessed and verified the underlying data. AK contributed to the study’s conception and supervised the data collection process in Uganda. All authors had full access to all the data in the study and accept responsibility for the decision to submit the manuscript for publication.

## Acknowledgements

This work was supported by the National Institutes of Health (grant numbers R01HL138728 [to E.A.K. and D.W.D.], T32 AI007291 and K23AI185268 [to J.S.]). The content is solely the responsibility of the authors and does not necessarily represent the official views of the National Institutes of Health.

