## supplementary appendix for "Performance of Universal and Stratified Computer-Aided Detection Thresholds for Chest X-Ray-Based Tuberculosis Screening: A Cross-Sectional Diagnostic Accuracy Study"

**Supplementary Appendix Table of Contents**

**Appendix A. Sample size and power calculation**

Of the 45,758 participants screened after the CAD threshold was lowered to 0.1, 284 participants with X-ray scores ≥0.1 tested positive on sputum Xpert. Assuming a TB prevalence of 0.1% among participants with X-ray scores <0.1, a total of 316 participants with Xpert-positive TB would be expected to contribute to the sensitivity estimate in the primary analysis. For the comparison of universal versus age- and sex-stratified CAD thresholds, for which empirical data showed correlation of 0.88, this sample size provides 80% power to detect a 3-percentage-point difference in sensitivity (e.g., 80% vs. 83%), with a two-sided alpha of 0.05 (calculated using the power.prop.ps function from the R package statpsych).

### Appendix B. Estimating Xpert positivity for participants with an X-ray score below 0.1

To estimate the prevalence of Xpert positivity among individuals with X-ray scores <0.1, we first considered national TB prevalence data as a benchmark. The 2014–2015 national TB prevalence survey in Uganda[1] estimated the prevalence of bacteriologically confirmed TB among individuals >15 years old at 401 per 100,000 population (0.4%). Given an approximately 80% (i.e., 77-80%) Xpert Ultra positivity rate among culture-positive individuals in other prevalence surveys,[2,3] a population with the same TB prevalence as 2014-2015 Ugandan average would have an Xpert-positive TB prevalence of approximately 0.3%. We expected the subset of study participants with X-ray scores <0.1 to have a substantially lower prevalence of TB than the overall population. Accordingly, we set 0.1% as our a priori estimate of TB prevalence among those with X-ray scores <0.1.

However, our study population as a whole appears to have a TB prevalence 2-3 times higher than this national average. Accordingly, we considered whether individuals with very low CAD scores in our study population might have a higher Xpert-TB prevalence (comparable to or higher than the national average prevalence). To explore this possibility, we considered data from another recent TB screening study[4] conducted in Uganda, in which Xpert Ultra testing was performed on all screening participants (regardless of symptoms, and without X-ray or other screening criteria). In that study, 9% of all individuals with an Xpert-positive or trace-positive sputum screening result, (19/219; 95% CI 5-13%) had CAD (qXR v4) scores less than 0.1 on a subsequent X-ray. We combined this estimate of low-CAD-score Xpert positivity with the estimates from the current study that 83% of participants had qXR scores >=0.1 and that 5% of these were Xpert-positive. Based on the simple equation below, we calculated that an Xpert positive prevalence of 0.1% among those with qXR score <0.1 would be needed in order for 9% of all people with positive Xpert to have qXR scores <0.1 (prevalence 0.05 – 0.15% across the uncertainty range of 5-13% above).

- **Equation for prevalence of Xpert positive TB among individuals with X-ray score<0.1**

*X=TB prevalence among people with score <0.1*

*Proportion of Xpert positive TB among people with CAD score<0.1*

$$= \frac{(\%\ of\ people\ with\ score\ < 0.1) * (TB\ prevalence\ among\ people\ with\ score\ < 0.1)}{(\%\ of\ people\ with\ score\ < 0.1) * (TB\ \%\ among\ people\ with\ score\ < 0.1) + (\%\ of\ people\ with\ score\ \geq 0.1) * (TB\ \%\ among\ people\ with\ score\ \geq 0.1)}$$

$$= \frac{(0.83 * X)}{(0.83 * X) + (0.17 * 0.05)} = 0.09$$

*∴ X=0.0010128 ≈0.1%*

As our a priori estimate of 0.1% Xpert positivity among participants with X-ray scores <0.1 was further supported by the available data, we used 0.1% as the assumed prevalence in the primary analysis. In sensitivity analyses, we explored a range from 0.05% (half of our estimate as a lower bound) up to 0.3% (i.e., assuming that the prevalence of TB among participants with lowest CAD scores in this study would be the same as among all adults in Uganda).

Lastly, as an alternative approach, we extrapolated probabilities of Xpert positivity across the range of X-ray scores from 0 to 0.1. To avoid underestimating the prevalence of TB among those with normal CXRs (which would lead to an over-estimation of CAD accuracy), we adopted intentionally conservative assumptions. First, we assumed that prevalence at a score of 0.09 was similar (i.e., 0.8%) to the average prevalence across the full 0.1-0.19 range (0.785%). From there, we modeled Xpert positivity as decreasing linearly, reaching 0.05% (still assumed to be greater than zero) at a score of 0. We also assumed that participants with X-ray scores under 0.1 had a similar probability of successfully providing sputum as those with X-ray scores between 0.1 and 0.2 (**Table S1**). This approach resulted in an overall Xpert positivity rate of 0.2% (84 out of the estimated 36,990 providing sputum) among participants with scores <0.1 (**Table S2**).

We interpreted this result – calculated from deliberately conservative assumptions – as supportive of our primary assumption of 0.1% positivity. The rather abrupt drop in aggregated Xpert positivity from the 0.1–0.2 to the 0.0–0.1 score bands (from 0.8% to 0.1%) is explained by the fact that half of the participants with a CAD score below 0.1 had extremely low scores of 0.00 or 0.01.

**Table S1. Number of participants at each X-ray score band with missing Xpert results**

| X-ray score | Total number of participants (n=52,835) | Participants not offered Xpert testing due to early study entry, n (%) | Participants declined or failed to provide a sputum sample for Xpert testing*, n(%) |
|---|---|---|---|
| 0.0 to 0.09 | 43,886 | N/A | N/A |
| 0.1 to 0.19 | 3,195 | 527 (16%) | 502 (16%) |
| 0.2 to 0.29 | 1,432 | 85 (6%) | 171 (12%) |
| 0.3 to 0.39 | 886 | 50 (6%) | 89 (10%) |
| 0.4 to 0.49 | 594 | 31 (5%) | 65 (11%) |
| 0.5 to 0.59 | 463 | N/A | 42 (9%) |
| 0.6 to 0.69 | 391 | N/A | 30 (8%) |
| 0.7 to 0.79 | 359 | N/A | 35 (10%) |
| 0.8 to 0.89 | 403 | N/A | 27 (7%) |
| 0.9 to 1.0 | 1,226 | N/A | 78 (6%) |

*This excludes participants not offered sputum testing due to early study entry.

**Table S2. Distribution of CAD scores among participants with scores below 0.1, with the estimated number of participants providing sputum samples and assumed Xpert positivity at each score.**

| X-ray score | No. of participants | Estimated No.* of participants providing a sputum sample | 0.1% TB prevalence among participants with X-ray scores <0.1 | |
|---|---|---|---|---|
| | | | Xpert positivity | No. of Xpert-positive participants |
| 0.00 | 3,333 | 2,809 | 0.05% | 1 |
| 0.01 | 20,007 | 16,863 | 0.13% | 22 |
| 0.02 | 8,368 | 7,053 | 0.22% | 15 |
| 0.03 | 4,258 | 3,589 | 0.30% | 11 |
| 0.04 | 2,515 | 2,120 | 0.38% | 8 |
| 0.05 | 1,750 | 1,475 | 0.47% | 7 |
| 0.06 | 1,286 | 1,084 | 0.55% | 6 |
| 0.07 | 963 | 812 | 0.63% | 5 |
| 0.08 | 797 | 672 | 0.72% | 5 |
| 0.09 | 609 | 513 | 0.80% | 4 |
| **0 to 0.09** | **43,886** | **36,990** | **0.23%** | **84** |

** Assumed to be 84% of participants with the same X-ray score*

### Appendix C. Methods to simulate screening populations and estimate accuracy outcomes in all participants and corresponding uncertainty

In order to estimate CAD accuracy outcomes and corresponding uncertainty among all participants, including those who were not asked to provide sputum due to their early study entry and those who failed or declined to provide a sputum sample, we first characterized the distribution of X-ray scores in intervals of width 0.1. Then we performed the following bootstrap simulation: Within each X-ray score interval, we estimated the number of participants who would have had valid Xpert results had all participants been asked to provide sputum. For X-ray scores ≥0.1, we then sampled that number of records with replacement from those with valid Xpert results in the same X-ray score interval. For scores <0.1, we randomly assigned an Xpert status based on an assumed Xpert-positive prevalence. This approach assumes that participants who were not asked to provide sputum had a comparable chance of testing positive on sputum Xpert (and similar chance of failing to provide sputum) as others with a similar CAD score. Finally, we combined the sampled records into a simulated cohort for calculating sensitivities and specificities of various Xpert testing thresholds, and we report confidence intervals as the 2.5$^{th}$ and 97.5$^{th}$ quantile across simulated cohorts

**Table S3: Characteristics of participants who received digital chest X-ray based TB screening, according to their Xpert status**

| | Xpert-negative participants (n=6,837) | Xpert-positive* participants (n=382) | p-value |
|---|---|---|---|
| **Age, median years (IQR)** | 52 (38, 66) | 39 (30, 51) | <0.001 |
| **Male, n (%)** | 3,554 (52.0%) | 289 (75.7%) | <0.001 |
| **Known HIV positive, n (%)** | 573 (8.4%) | 25 (6.5%) | 0.205 |
| **Prior TB, n (%)** | 355 (5.2%) | 47 (12.3%) | 0.001 |
| **Prior household TB exposure, n (%)** | 105 (1.5%) | 13 (3.4%) | 0.005 |
| **Current smoking**, n/N (%)** | 166/1,972 (8.4%) | 13/86 (15.1%) | 0.031 |
| **Symptom within 30 days** | | | |
| Cough, n (%) | 2,931 (42.9%) | 284 (74.3%) | <0.001 |
| Cough ≥ 2 weeks, n (%) | 1,958 (28.6%) | 245 (64.1%) | <0.001 |
| Fever, n (%) | 1,535 (22.5%) | 117 (30.6%) | <0.001 |
| Night sweats, n (%) | 373 (5.5%) | 60 (15.7%) | <0.001 |
| Weight loss, n (%) | 235 (3.4%) | 46 (12.0%) | <0.001 |
| Any TB symptom[†], n (%) | 3,640 (53.2%) | 308 (80.6%) | <0.001 |
| **X-ray score, median (IQR)** | 0.31 (0.17, 0.64) | 0.97 (0.82, 0.99) | <0.001 |

* Includes Ultra trace-positive results

** Only asked for participants enrolled after November 11, 2023

[†] Any cough, fever, night sweats, or weight loss

**Table S4: Characteristics of participants with X-ray score <0.5 with valid Xpert results**

| | Xpert-negative participants (n=4,534) | Xpert-positive* participants (n=55) | p-value |
|---|---|---|---|
| **Age, median years (IQR)** | 52 (38, 65) | 41 (32, 56) | 0.005 |
| **Male, n (%)** | 2,211 (48.8%) | 39 (70.9%) | 0.001 |
| **Known HIV positive, n (%)** | 330 (7.3%) | 2 (3.6%) | 0.300 |
| **Prior TB, n (%)** | 66 (1.5%) | 2 (3.6%) | <0.001 |
| **Prior household TB exposure, n (%)** | 66 (1.5%) | 2 (3.6%) | 0.183 |
| **Current smoking**, n/N (%)** | 101/1,322 (7.6%) | 1/9 (11.1%) | 0.696 |
| **Symptom within 30 days** | | | |
| Cough, n (%) | 1664 (36.7%) | 33 (60.0%) | <0.001 |
| Cough ≥ 2 weeks, n (%) | 1015 (22.4%) | 27 (49.1%) | <0.001 |
| Fever, n (%) | 968 (21.3%) | 11 (20.0%) | 0.808 |
| Night sweats, n (%) | 209 (4.6%) | 7 (12.7%) | 0.005 |
| Weight loss, n (%) | 113 (2.5%) | 4 (7.3%) | 0.025 |
| Any TB symptom†, n (%) | 2166 (47.8%) | 37 (67.3%) | 0.004 |
| **X-ray score, median (IQR)** | 0.20 (0.14, 0.31) | 0.29 (0.18, 0.38) | <0.001 |

* Includes Ultra trace-positive results

** Only asked for participants enrolled after November 11, 2023

† Any cough, fever, night sweats, or weight loss

**Table S5: Proportion of participants with abnormal X-ray and positive sputum Xpert Ultra results, by sex and age group**

| | Male (n=23.586) | Female (n=29,249) | < 40 years old (n=28,227) | ≥ 40 years old (n=24,607) | All (n=52,835) |
|---|---|---|---|---|---|
| **X-ray score ≥0.1** | 4,726 (20.2%) | 4,223 (14.4%) | 2552 (9.0%) | 6,396 (26.0%) | 8,949 (16.9%) |
| **X-ray score ≥0.5** | 1,705 (7.2%) | 1,137 (3.9%) | 816 (2.9%) | 2,026 (8.2%) | 2,842 (5.4%) |
| **Positive* Xpert Ultra** | 289 (1.2%) | 93 (0.3%) | 193 (0.7%) | 189 (0.8%) | 382 (0.7%) |

*Includes Ultra trace-positive results

**Figure S1. Correlation between X-ray scores and semiquantitative Xpert results, among participants who tested positive on sputum Xpert**

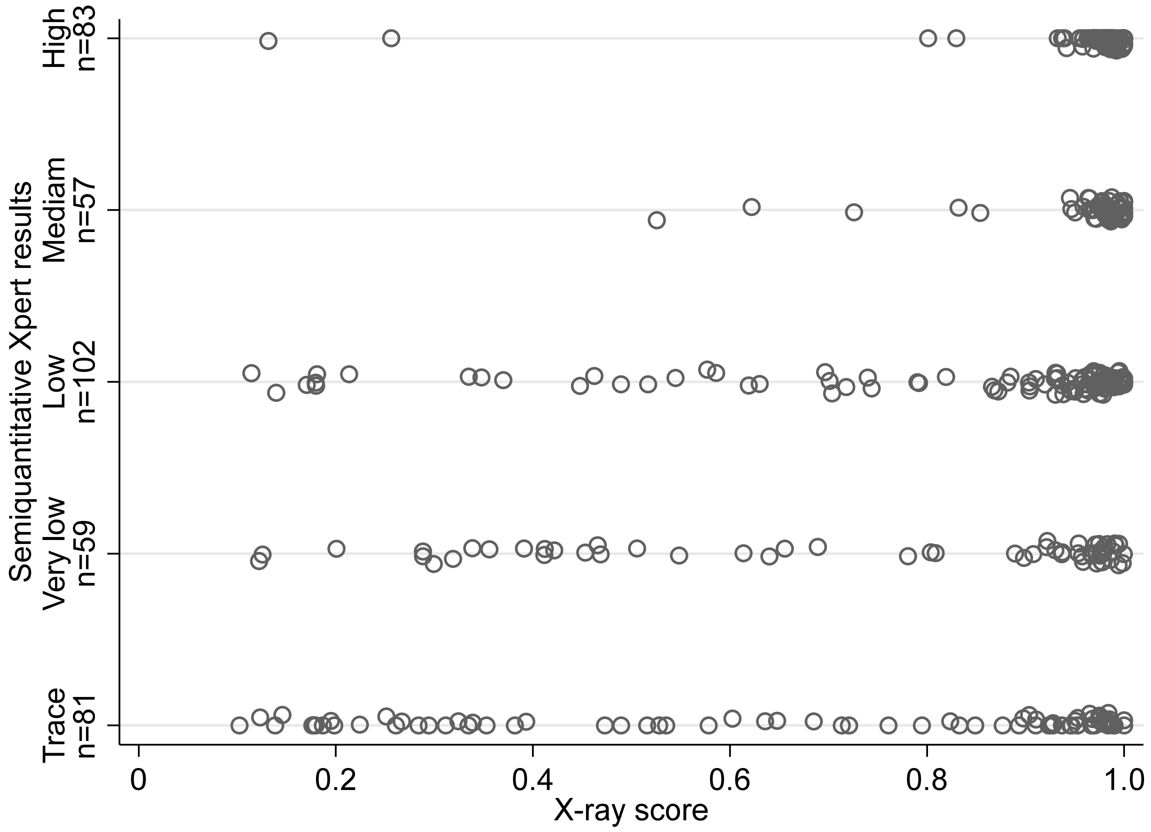

**Table S6: Sensitivities and specificities of digital X-ray with computer-aided detection software (qXR) for tuberculosis screening, using fixed thresholds**

| | Assumed TB prevalence, score <0.1 | X-ray score threshold | | | | | | | | |
|---|---|---|---|---|---|---|---|---|---|---|
| | | ≥0.1 | ≥0.2 | ≥0.3 | ≥0.4 | ≥0.5 | ≥0.6 | ≥0.7 | ≥0.8 | ≥0.9 |
| **Sensitivity, %<br>95% CI** | **0.05%** | 94.7<br>91.5–96.7) | 89.0<br>85.0–92.1 | 87.3<br>83.1–90.6 | 85.0<br>80.5–88.6 | 83.3<br>78.7–87.1 | 80.7<br>75.8–84.7 | 77.3<br>72.3–81.7 | 75.0<br>69.8–79.6 | 70.3<br>64.9–75.2 |
| **Specificity, %<br>95% CI** | **0.05%** | 83.6<br>83.2–83.9) | 89.2<br>88.8–89.5 | 92.0<br>91.7–92.2 | 93.7<br>93.4–93.9 | 94.8<br>94.6–95.0 | 95.7<br>95.5–95.9 | 96.5<br>96.3–96.7 | 97.2<br>97.1–97.4 | 98.0<br>97.9–98.2 |
| **AUC (95% CI)** | **0.05%** | 0.979 (0.975-0.983) | | | | | | | | |
| **Sensitivity, %<br>95% CI** | **0.1%** | 89.9<br>86.1–92.7 | 84.5<br>80.1–88.1 | 82.9<br>78.4–86.7 | 80.7<br>76.0–84.7 | 79.1<br>74.3–83.2 | 76.6<br>71.6–80.9 | 73.4<br>68.3–78.0 | 71.2<br>66.0–75.9 | 66.8<br>61.4–71.7 |
| **Specificity, %<br>95% CI** | **0.1%** | 83.6<br>83.2–83.9 | 89.2<br>88.8–89.5 | 92.0<br>91.7–92.2 | 93.7<br>93.4–93.9 | 94.8<br>94.6–95.0 | 95.7<br>95.5–95.9 | 96.5<br>96.3–96.7 | 97.2<br>97.1–97.4 | 98.0<br>97.9–98.2 |
| **AUC (95% CI)** | **0.1%** | 0.920 (0.898-0.941) | | | | | | | | |
| **Sensitivity, %<br>95% CI** | **0.2%** | 81.6<br>77.2–85.3 | 76.7<br>72.0–80.9 | 75.3<br>70.5–79.5 | 73.3<br>68.4–77.7 | 71.8<br>66.9–76.3 | 69.5<br>64.5–74.1 | 66.7<br>61.6–71.4 | 64.7<br>59.5–69.5 | 60.6<br>55.4–65.6 |
| **Specificity, %<br>95% CI** | **0.2%** | 83.6<br>83.2–83.9 | 89.1<br>88.8–89.5 | 92.0<br>91.7–92.2 | 93.7<br>93.4–93.9 | 94.8<br>94.6–95.0 | 95.7<br>95.5–95.9 | 96.5<br>96.3–96.7 | 97.2<br>97.1–97.4 | 98.0<br>97.9–98.2 |
| **AUC (95% CI)** | **0.2%** | 0.877 (0.853-0.902) | | | | | | | | |
| **Sensitivity, %<br>95% CI** | **0.3%** | 74.5<br>69.9–78.7 | 70.1<br>65.3–74.5 | 68.8<br>63.9–73.2 | 66.9<br>62.1–71.5 | 65.6<br>60.7–70.2 | 63.5<br>58.6–68.2 | 60.9<br>55.9–65.7 | 59.1<br>54.1–63.9 | 55.4<br>50.4–60.3 |
| **Specificity, %<br>95% CI** | **0.3%** | 83.5<br>83.2–83.9 | 89.1<br>88.8–89.4 | 91.9<br>91.7–92.2 | 93.7<br>93.4–93.9 | 94.8<br>94.6–95.0 | 95.7<br>95.5–95.9 | 96.5<br>96.3–96.7 | 97.2<br>97.1–97.4 | 98.0<br>97.9–98.2 |
| **AUC (95% CI)** | **0.3%** | 0.834 (0.806-862) | | | | | | | | |

**Figure S2. Sensitivities and specificities of digital chest X-ray with computer-aided detection software (qXR) for tuberculosis screening, by screening location.** Plots below illustrate the sensitivity and specificity of the qXR v3 software in detecting sputum Xpert-positive tuberculosis across eight regions surrounding Kampala. Dots represent estimates and bars indicate 95% confidence intervals calculated by the Wilson score method. Estimates among all screening participants capable of providing sputum were calculated assuming a 0.1% prevalence of sputum Xpert-positive TB among individuals with X-ray scores <0.1. In the "original" analysis, Xpert-positive TB cases with scores <0.1 were randomly assigned across the eight regions. In the "adjusted" analysis, the prevalence of TB among individuals with X-ray scores <0.1 in each region was assumed to be proportionate to the prevalence among individuals with scores ≥0.1 in the same region.

#### a) Sensitivity, threshold of 0.1

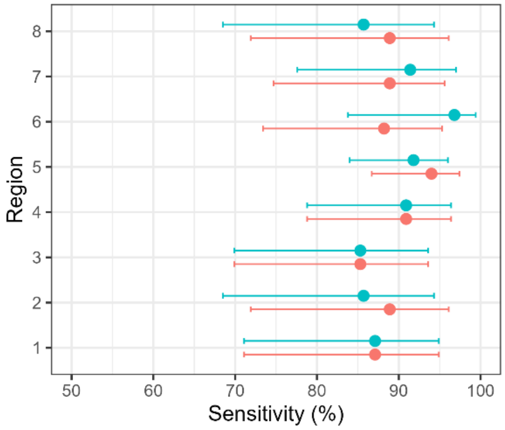

#### b) Specificity, threshold of 0.1

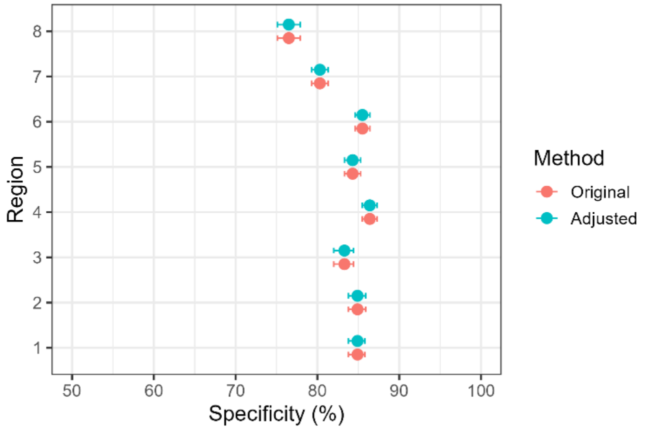

#### c) Sensitivity, threshold of 0.5

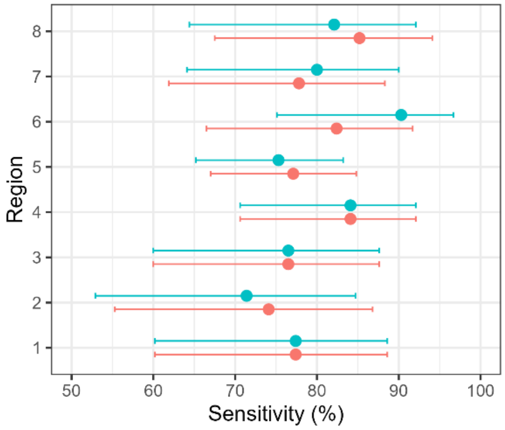

#### d) Specificity, threshold of 0.5

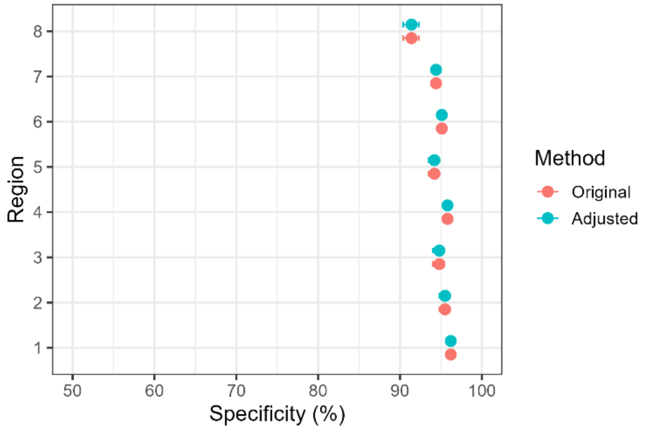

**Figure S3. Receiver operating characteristic curves illustrating diagnostic performance of computer-aided detection software (qXR version 3) for sputum Xpert-positive TB during community-based TB screening in Uganda, among all participants eligible for screening.** The colors of the ROC curves represent different assumptions about the prevalence of TB among people with an X-ray score <0.1 who did not qualify for sputum Xpert testing (red line: 0% prevalence; green line: 0.1% prevalence; blue line: 0.3% prevalence). Bands display 95% confidence intervals.

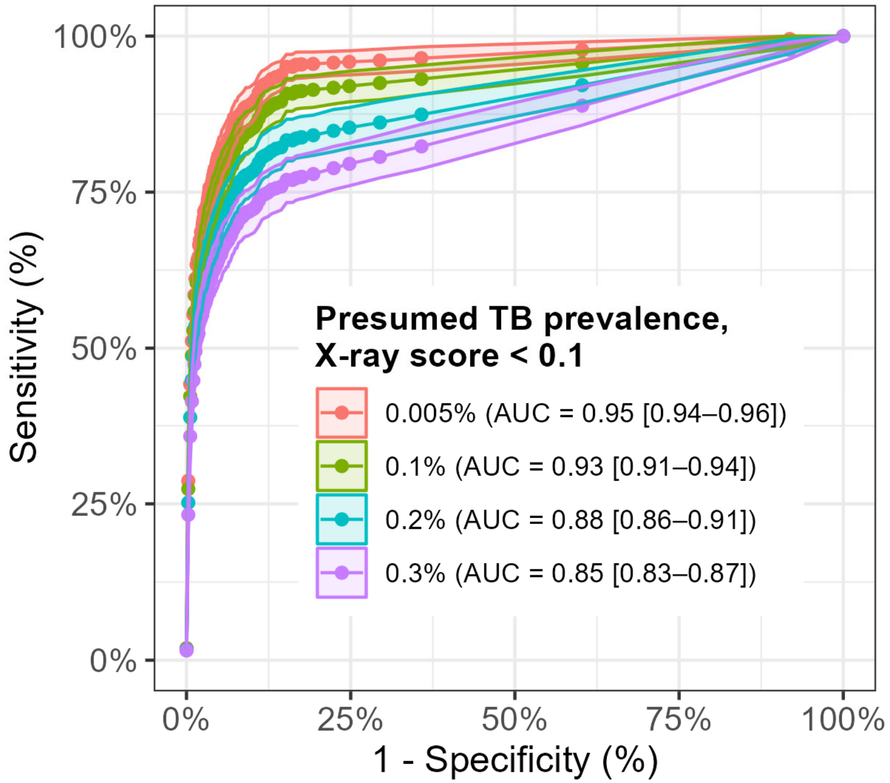

**Table S7: Sensitivity and specificity of age- and sex- stratified qXR thresholds for detecting Xpert-positive tuberculosis, compared with universal thresholds of matching specificity, considering trace-positive results as negative results**

| | Assumed TB prevalence when X-ray score <0.1 | Age- and sex-stratified threshold* (15-39y M: ≥0.11; ≥40y M: ≥0.25; 15-39y F: ≥0.28; ≥40y F: 0.44) | Universal threshold (≥0.26 for all†) | Age- and sex-stratified threshold* (15-39y M: ≥0.47; ≥40y M: ≥0.53; 15-39y F: ≥0.52; ≥40y F: 0.89) | Universal threshold (≥0.64 for all†) |
|---|---|---|---|---|---|
| *Sensitivity* | 0.05% | 90.7 | 89.5 | 84.2 | 82.6 |
| *95% CI* | | 86.4-93.7 | 85.0-92.7 | 79.1-88.2 | 77.4-86.8 |
| *Specificity* | 0.05% | 90.9 | 90.9 | 96.0 | 96.0 |
| *95% CI* | | 90.6-91.2 | 90.6-91.2 | 95.8-96.2 | 95.8-96.2 |
| *Sensitivity* | 0.1% | 85.2 | 84.0 | 79.1 | 77.6 |
| *95% CI* | | 80.4-89.0 | 79.1-88.0 | 73.8-83.6 | 72.1-82.2 |
| *Specificity* | 0.1% | 90.9 | 90.9 | 96.0 | 96.0 |
| *95% CI* | | 90.6-91.2 | 90.6-91.2 | 95.8-96.2 | 95.8-96.2 |
| *Sensitivity* | 0.2% | 75.9 | 74.9 | 70.5 | 69.2 |
| *95% CI* | | 70.7-80.5 | 69.7-79.5 | 65.1-75.4 | 63.7-74.1 |
| *Specificity* | 0.2% | 90.9 | 90.9 | 96.0 | 96.0 |
| *95% CI* | | 90.6-91.2 | 90.6-91.2 | 95.8-96.2 | 95.8-96.2 |
| *Sensitivity* | 0.3% | 68.3 | 67.4 | 63.4 | 62.2 |
| *95% CI* | | 63.1-73.1 | 62.1-72.2 | 58.1-68.4 | 56.8-67.3 |
| *Specificity* | 0.3% | 90.9 | 90.9 | 96.0 | 96.0 |
| *95% CI* | | 90.6-91.2 | 90.6-91.2 | 95.8-96.2 | 95.8-96.2 |

* Correspond to the estimated scores at which approximately 1% (left columns) or 2% (right columns) of participants in each subgroup would test positive.

† Chosen to match the 90.9% (left columns) or 96.0% (right columns) aggregate specificity of age- and sex-stratified thresholds.

**Table S8: Sensitivity and specificity of age or sex-stratified qXR thresholds for detecting Xpert-positive tuberculosis, compared with universal thresholds of matching specificity, considering trace-positive results as negative results**

| | Assumed TB prevalence when X-ray score <0.1 | Age-stratified threshold* (15-39y: ≥0.19; ≥40y: ≥0.38) | Universal threshold (≥0.31 for all†) | Age-stratified threshold* (15-39y: ≥0.47; ≥40y: ≥0.62) | Universal threshold (≥0.59 for all†) | Sex-stratified threshold* (Male: ≥0.08; Female: ≥0.43) | Universal threshold (≥0.16 for all†) | Sex-stratified threshold* (Male: ≥0.52; Female: ≥0.81) | Universal threshold (≥0.64 for all†) |
|---|---|---|---|---|---|---|---|---|---|
| *Sensitivity* | 0.05% | 88.7 | 88.7 | 84.6 | 84.2 | 91.9 | 91.5 | 83.0 | 82.6 |
| *95% CI* | | 84.1-92.0 | 84.1-92.0 | 79.6-88.6 | 79.1-88.2 | 87.8-94.7 | 87.4-94.4 | 77.8-87.2 | 77.4-86.8 |
| *Specificity* | 0.05% | 92.1 | 92.1 | 95.6 | 95.6 | 87.5 | 87.4 | 96.0 | 96.0 |
| *95% CI* | | 91.9-92.4 | 91.8-92.13 | 95.3-95.8 | 95.4-95.8 | 87.2-87.8 | 87.1-87.8 | 95.8-96.2 | 95.8-96.2 |
| *Sensitivity* | 0.1% | 83.3 | 83.3 | 79.5 | 79.1 | 86.3 | 85.9 | 77.9 | 77.6 |
| *95% CI* | | 78.3-87.3 | 78.3-87.3 | 74.2-83.9 | 73.8-83.6 | 81.6-89.9 | 81.2-89.6 | 72.6-82.5 | 72.1-82.2 |
| *Specificity* | 0.1% | 92.1 | 92.0 | 95.6 | 95.6 | 87.5 | 87.4 | 96.0 | 96.0 |
| *95% CI* | | 91.9-92.4 | 91.8-92.3 | 95.3-95.8 | 95.4-95.8 | 87.2-87.8 | 87.1-87.8 | 95.8-96.2 | 95.8-96.2 |
| *Sensitivity* | 0.2% | 74.2 | 74.2 | 70.8 | 70.5 | 77.3 | 76.6 | 69.5 | 69.2 |
| *95% CI* | | 69.0-78.9 | 69.0-78.9 | 65.4-75.7 | 65.1-75.4 | 72.2-81.7 | 71.5-81.1 | 64.0-74.5 | 63.7-74.1 |
| *Specificity* | 0.2% | 92.1 | 92.0 | 95.6 | 95.6 | 87.5 | 87.4 | 96.0 | 96.0 |
| *95% CI* | | 91.8-92.4 | 91.8-92.3 | 95.3-95.8 | 95.4-95.8 | 87.2-87.8 | 87.1-87.7 | 95.8-96.2 | 95.8-96.2 |
| *Sensitivity* | 0.3% | 66.8 | 66.8 | 63.7 | 63.4 | 69.5 | 68.9 | 62.5 | 62.2 |
| *95% CI* | | 61.5-71.6 | 61.5-71.6 | 58.4-68.7 | 58.1-68.4 | 64.3-74.2 | 63.7-73.7 | 57.1-67.6 | 56.8-67.3 |
| *Specificity* | 0.3% | 92.1 | 92.0 | 95.6 | 95.6 | 87.5 | 87.4 | 96.0 | 96.0 |
| *95% CI* | | 91.8-92.4 | 91.8-92.3 | 95.3-95.8 | 95.4-95.8 | 87.1-87.8 | 87.1-87.7 | 95.8-96.2 | 95.8-96.2 |

* Correspond to the estimated scores at which approximately 1% (lower thresholds) or 2% (higher threshold) of participants in each subgroup would test positive.

† Chosen to match the aggregate specificity of corresponding stratified thresholds.

**Table S9: Sensitivity and specificity of age- and sex- stratified qXR thresholds for detecting Xpert-positive tuberculosis, compared with universal thresholds of matching specificity, among all participants eligible for screening**

| | Assumed TB prevalence when X-ray score <0.1 | Age- and sex-stratified threshold* (15-39y M: ≥0.11; ≥40y M: ≥0.25; 15-39y F: ≥0.28; ≥40y F: 0.44) | Universal threshold (≥0.26 for all†) | Age- and sex-stratified threshold* (15-39y M: ≥0.47; ≥40y M: ≥0.53; 15-39y F: ≥0.52; ≥40y F: 0.89) | Universal threshold (≥0.64 for all†) |
|---|---|---|---|---|---|
| *Sensitivity*<br>*95% CI* | 0.05% | 89.7<br>86.5-92.7 | 88.4<br>85.3-91.3 | 77.0<br>73.0-80.9 | 76.0<br>72.0-80.1 |
| *Specificity*<br>*95% CI* | 0.05% | 91.1<br>91.0-91.3 | 91.2<br>91.1-91.3 | 96.2<br>96.1-96.3 | 96.2<br>96.1-96.3 |
| *Sensitivity*<br>*95% CI* | 0.1% | 85.6<br>82.1-88.9 | 84.4<br>80.7-87.5 | 73.4<br>69.4-77.5 | 72.4<br>68.5-76.5 |
| *Specificity*<br>*95% CI* | 0.1% | 91.1<br>91.0-91.3 | 91.2<br>91.1-91.3 | 96.2<br>96.1-96.3 | 96.2<br>96.1-96.3 |
| *Sensitivity*<br>*95% CI* | 0.2% | 78.6<br>75.1-82.3 | 77.5<br>73.6-81.2 | 67.6<br>63.2-71.6 | 66.7<br>62.4-70.7 |
| *Specificity*<br>*95% CI* | 0.2% | 91.1<br>91.0-91.2 | 91.2<br>91.1-91.3 | 96.2<br>96.1-96.3 | 96.2<br>96.1-96.3 |
| *Sensitivity*<br>*95% CI* | 0.3% | 72.6<br>68.9-76.3 | 71.5<br>67.7-75.1 | 62.3<br>58.3-66.0 | 61.6<br>57.5-65.2 |
| *Specificity*<br>*95% CI* | 0.3% | 91.1<br>90.9-91.2 | 91.2<br>91.1-91.3 | 96.2<br>96.1-96.3 | 96.2<br>96.1-96.3 |

* Correspond to the estimated scores at which approximately 1% (left columns) or 2% (right columns) of participants in each subgroup would test positive.

† Chosen to match the 91.1% (left columns) or 96.2% (right columns) aggregate specificity of age- and sex-stratified thresholds.

**Table S10: Sensitivity and specificity of age or sex-stratified qXR thresholds for detecting Xpert-positive tuberculosis, compared with universal thresholds of matching specificity, among all participants eligible for screening**

| | Assumed TB prevalence when X-ray score <0.1 | Age-stratified threshold* (15-39y: ≥0.19; ≥40y: ≥0.38) | Universal threshold (≥0.31 for all†) | Age-stratified threshold* (15-39y: ≥0.47; ≥40y: ≥0.62) | Universal threshold (≥0.58 for all†) | Sex-stratified threshold* (Male: ≥0.08; Female: ≥0.43) | Universal threshold (≥0.18 for all†) | Sex-stratified threshold* (Male: ≥0.52; Female: ≥0.81) | Universal threshold (≥0.64 for all†) |
|---|---|---|---|---|---|---|---|---|---|
| *Sensitivity* | 0.05% | 87.0 | 86.3 | 77.7 | 77.7 | 91.9 | 91.9 | 76.9 | 76.0 |
| *95% CI* | | 83.5-90.1 | 82.9-89.6 | 73.6-81.7 | 73.8-81.6 | 88.9-94.5 | 89.1-94.4 | 72.9-80.8 | 72.0-80.1 |
| *Specificity* | 0.05% | 92.4 | 92.3 | 95.7 | 95.7 | 88.5 | 88.5 | 96.2 | 96.2 |
| *95% CI* | | 92.3-92.5 | 92.3-92.4 | 95.7-95.8 | 95.6-95.8 | 87.9-89.0 | 88.4-88.6 | 96.1-96.3 | 96.1-96.3 |
| *Sensitivity* | 0.1% | 82.9 | 82.4 | 74.0 | 74.1 | 87.7 | 87.7 | 73.4 | 72.4 |
| *95% CI* | | 79.1-86.2 | 78.7-85.9 | 70.0-78.0 | 70.0-78.0 | 84.4-90.7 | 84.5-90.6 | 69.1-77.5 | 68.5-76.5 |
| *Specificity* | 0.1% | 92.4 | 92.3 | 95.7 | 95.7 | 88.5 | 88.5 | 96.2 | 96.2 |
| *95% CI* | | 92.3-92.5 | 92.3-92.4 | 95.7-95.8 | 95.6-95.8 | 87.9-89.0 | 88.4-88.6 | 96.1-96.3 | 96.1-96.3 |
| *Sensitivity* | 0.2% | 76.3 | 75.7 | 68.2 | 68.1 | 80.5 | 80.5 | 67.5 | 66.7 |
| *95% CI* | | 72.3-80.0 | 71.7-79.5 | 64.0-72.4 | 64.0-72.3 | 77.1-84.1 | 76.9-84.1 | 63.2-71.7 | 62.4-70.7 |
| *Specificity* | 0.2% | 92.4 | 92.3 | 95.7 | 95.7 | 88.5 | 88.5 | 96.2 | 96.2 |
| *95% CI* | | 92.3-92.5 | 92.3-92.4 | 95.7-95.8 | 95.6-95.8 | 87.9-89.0 | 88.4-88.6 | 96.1-96.3 | 96.1-96.3 |
| *Sensitivity* | 0.3% | 70.3 | 69.8 | 62.9 | 62.9 | 74.4 | 74.4 | 62.3 | 61.6 |
| *95% CI* | | 66.5-74.1 | 66.0-73.4 | 58.7-66.6 | 58.9-66.5 | 70.6-78.0 | 70.6-78.0 | 58.1-66.1 | 57.5-65.2 |
| *Specificity* | 0.3% | 92.4 | 92.3 | 95.7 | 95.7 | 88.5 | 88.5 | 96.2 | 96.2 |
| *95% CI* | | 92.3-92.5 | 92.3-92.4 | 95.7-95.8 | 95.6-95.8 | 87.9-89.0 | 88.4-88.6 | 96.1-96.3 | 96.1-96.3 |

* Correspond to the estimated scores at which approximately 1% (lower thresholds) or 2% (higher threshold) of participants in each subgroup would test positive.

† Chosen to match the aggregate specificity of corresponding stratified thresholds.

**Table S11: Sensitivity and specificity of age or sex-stratified qXR thresholds for detecting Xpert-positive tuberculosis, compared with universal thresholds of matching specificity**

| | Assumed TB prevalence when X-ray score <0.1 | Age-stratified threshold* (15-39y: ≥0.19; ≥40y: ≥0.38) | Universal threshold (≥0.31 for all†) | Age-stratified threshold* (15-39y: ≥0.47; ≥40y: ≥0.62) | Universal threshold (≥0.58 for all†) | Sex-stratified threshold* (Male: ≥0.08; Female: ≥0.43) | Universal threshold (≥0.16 for all†) | Sex-stratified threshold* (Male: ≥0.52; Female: ≥0.81) | Universal threshold (≥0.64 for all†) |
|---|---|---|---|---|---|---|---|---|---|
| *Sensitivity* | 0.05% | 87.3 | 86.7 | 80.7 | 81.3 | 91.7 | 91.7 | 80.0 | 79.0 |
| *95% CI* | | 83.1-90.6 | 82.4-90.1 | 75.8-84.7 | 76.5-85.3 | 88.0-94.3 | 88.0-94.3 | 75.1-84.1 | 74.0-83.2 |
| *Specificity* | 0.05% | 92.2 | 92.1 | 95.6 | 95.6 | 87.6 | 87.5 | 96.1 | 96.1 |
| *95% CI* | | 92.0-92.5 | 91.9-92.4 | 95.4-95.8 | 95.4-95.8 | 87.3-87.9 | 87.2-87.9 | 95.9-96.3 | 59.9-96.3 |
| *Sensitivity* | 0.1% | 82.9 | 82.3 | 76.6 | 77.2 | 87.0 | 87.0 | 75.9 | 75.0 |
| *95% CI* | | 78.4-86.7 | 77.7-86.1 | 71.6-80.9 | 72.3-81.5 | 82.9-90.3 | 82.9-90.3 | 70.9-80.3 | 69.9-79.5 |
| *Specificity* | 0.1% | 92.2 | 92.1 | 95.6 | 95.6 | 87.6 | 87.5 | 96.1 | 96.1 |
| *95% CI* | | 92.0-92.5 | 91.9-92.4 | 95.4-95.8 | 95.4-95.8 | 87.3-87.9 | 87.2-87.9 | 95.9-96.3 | 95.9-96.3 |
| *Sensitivity* | 0.2% | 75.3 | 74.7 | 69.5 | 70.1 | 79.6 | 79.0 | 69.0 | 68.1 |
| *95% CI* | | 70.5-79.5 | 69.9-79.0 | 64.5-74.1 | 65.1-74.7 | 75.1-83.5 | 74.4-83.0 | 63.9-73.6 | 63.0-72.8 |
| *Specificity* | 0.2% | 92.2 | 92.1 | 95.6 | 95.6 | 87.6 | 87.5 | 96.1 | 96.1 |
| *95% CI* | | 91.9-92.5 | 91.9-92.4 | 95.4-95.8 | 95.4-95.8 | 87.3-87.9 | 87.2-87.9 | 95.9-96.3 | 95.9-96.3 |
| *Sensitivity* | 0.3% | 68.8 | 68.2 | 63.5 | 64.0 | 72.4 | 72.2 | 63.0 | 62.2 |
| *95% CI* | | 63.9-73.2 | 63.4-72.7 | 58.6-68.2 | 59.1-68.7 | 67.7-76.7 | 67.5-76.4 | 58.0-67.7 | 57.2-66.9 |
| *Specificity* | 0.3% | 92.2 | 92.1 | 95.6 | 95.6 | 87.6 | 87.5 | 96.1 | 96.1 |
| *95% CI* | | 91.9-92.5 | 91.9-92.4 | 95.4-95.8 | 95.4-95.8 | 87.3-87.9 | 87.2-87.8 | 95.9-96.3 | 95.9-96.3 |

* Correspond to the estimated scores at which approximately 1% (lower thresholds) or 2% (higher threshold) of participants in each subgroup would test positive.

† Chosen to match the aggregate specificity of corresponding stratified thresholds.

**Figure S4. Patient flow diagram illustrating the number of participants at each step and comparing outcomes using age- and sex-stratified thresholds selected to yield 2% Xpert positivity, and a universal threshold (≥0.64) matched for specificity**

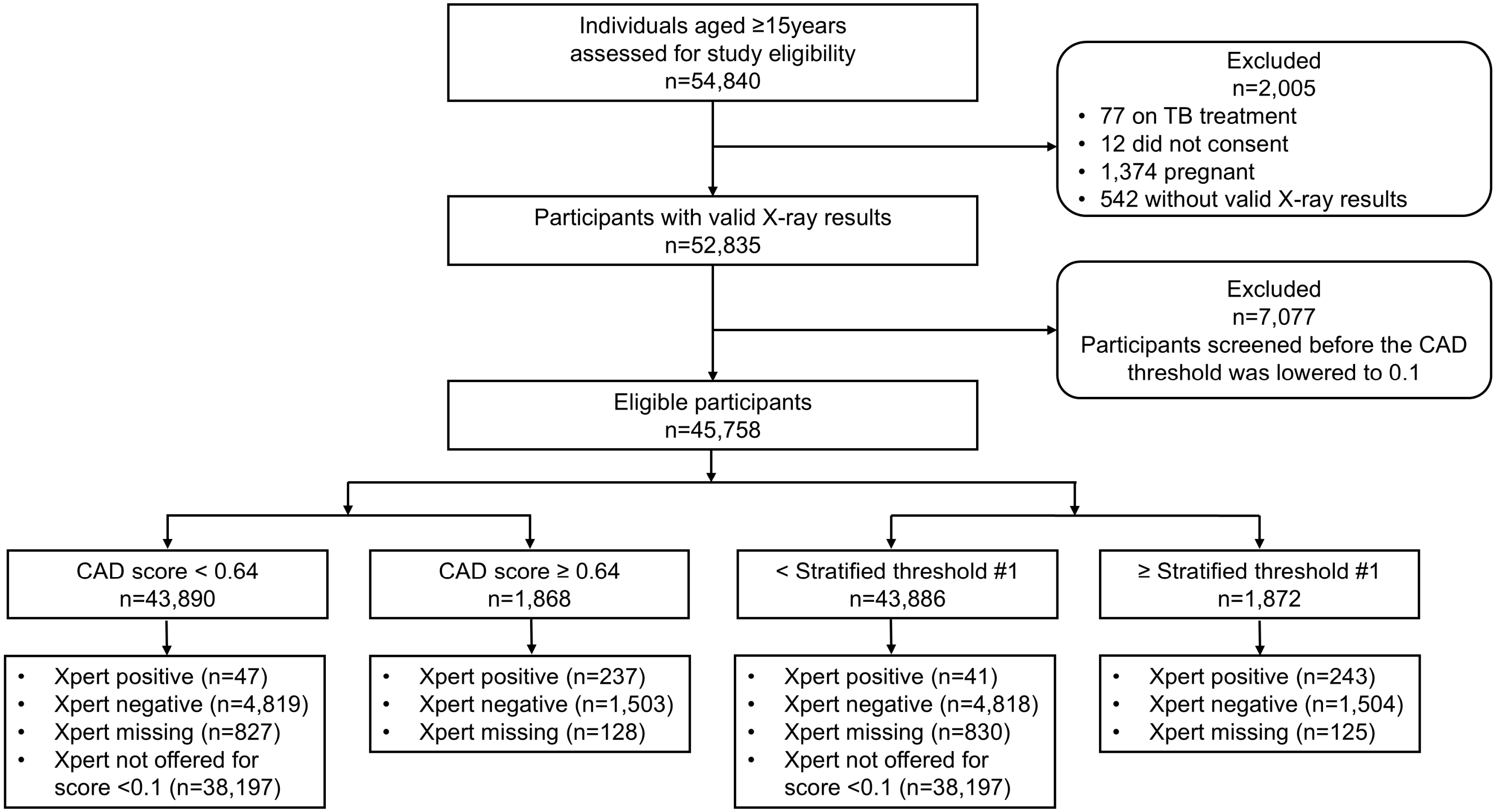

**Figure S5. Patient flow diagram illustrating the number of participants at each step and comparing outcomes using age- and sex-stratified thresholds selected to yield 1% Xpert positivity, and a universal threshold (≥0.26) matched for specificity**

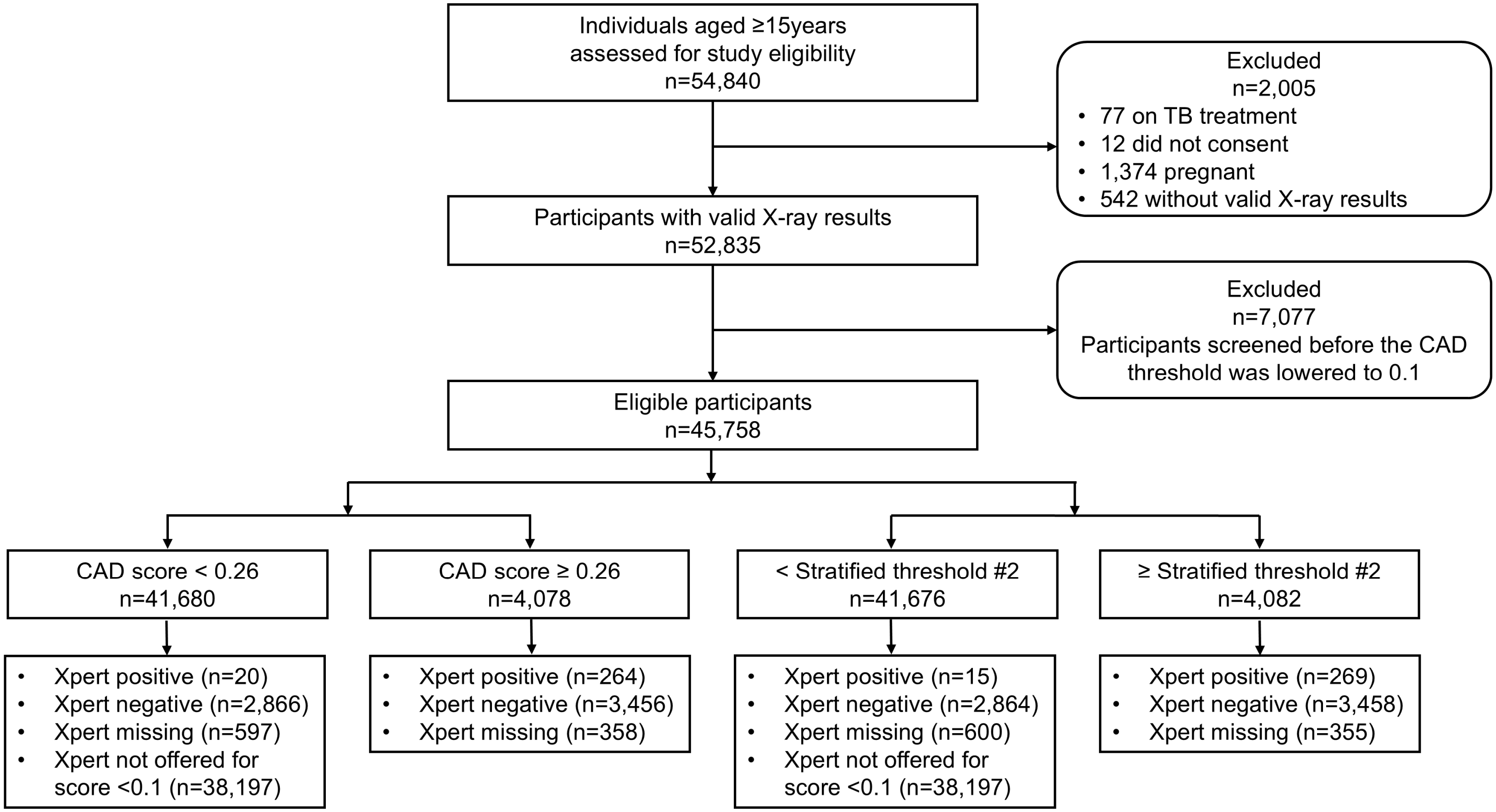

**Table S12. Subgroup thresholds required to achieve ≥80% or ≥90% sensitivity, or ≥70% specificity within each subgroup, assuming a 0.1% prevalence of Xpert-positive tuberculosis among individuals with an X-ray score below 0.1**

| Subgroup | Target | Threshold | Subgroup sensitivity | Subgroup specificity |
|---|---|---|---|---|
| Male | Sensitivity ≥ 80% | 0.62 | 80.7% | 95.2% |
| | Sensitivity ≥ 90% | 0.18 | 90.6% | 87.6% |
| | Specificity ≥ 70% | 0.05 | 95.1% | 70.6% |
| Female | Sensitivity ≥ 80% | 0.09 | 80.6% | 86.9% |
| | Sensitivity ≥ 90% | 0.01 | 96.8% | 9.9% |
| | Specificity ≥ 70% | 0.04 | 82.8% | 75.2% |
| < 40 years old | Sensitivity ≥ 80% | 0.47 | 80.0% | 97.9% |
| | Sensitivity ≥ 90% | 0.03 | 90.0% | 75.0% |
| | Specificity ≥ 70% | 0.03 | 90.0% | 75.0% |
| ≥ 40 years old | Sensitivity ≥ 80% | 0.39 | 80.3% | 90.8% |
| | Sensitivity ≥ 90% | 0.07 | 91.1% | 71.2% |
| | Specificity ≥ 70% | 0.07 | 91.1% | 71.2% |
